# Prevalence of and factors associated with malnutrition among women receiving PMTCT care at public hospitals in Addis Ababa: A cross-sectional study

**DOI:** 10.64898/2025.12.30.25343220

**Authors:** Fitsum Zekarias Mohammed, Senaite Abebe, Solomon Hailemeskel

## Abstract

**Background:** HIV and malnutrition act synergistically to weaken the immune system, increasing susceptibility to opportunistic infections, morbidity, and mortality. HIV destroys the body ability to fight infections, while malnutrition hinders recovery, accelerating the progression of AIDS related illnesses. This combination also undermines the adherence to and effectiveness of antiretroviral therapy (ART), particularly in resource limited settings. Accordingly, this study assessed the prevalence of malnutrition and associated factors among women attending prevention of mother to child transmission programs in public hospitals in Addis Ababa, Ethiopia, in 2024.

**Methods:** A cross sectional study was undertaken from April 1st to 20th, 2024. The study enrolled 193 women receiving PMTCT care at six public hospitals in Addis Ababa, selected through a simple random sampling method. Data were collected via face to face interviews using a standardized, structured questionnaire. Additionally, participants mid upper arm circumference (MUAC), weight, and height were measured to assess nutritional status. Data analysis was performed using SPSS version 26. Binary logistic regression was employed to examine the strength of associations, with results expressed as odds ratios alongside 95% confidence intervals. A p value score of 0.05 was considered statistically significant.

**Results:** The overall prevalence of malnutrition was 29% (19% undernutrition, 8% overweight, 2% obesity). In the adjusted analysis, younger age (25 to 34 years) was protective (AOR: 0.25), whereas experiencing eating problems (AOR: 13.70) and gastrointestinal symptoms (AOR: 3.52), immunosuppression (AOR: 8.13), anemia (AOR: 5.03), low meal frequency (AOR: 4.12), poor adherence (AOR: 3.60) were significant risk factors.

**Conclusion:** The prevalence of malnutrition among women receiving PMTCT at public hospitals in Addis Ababa was high. Moving forward, integrating routine nutritional screening and evidence based supportive interventions, including dietary support for women with low meal frequency and targeted management for those with anemia or low CD4 counts, should be a priority to improve comprehensive care for this vulnerable population.

## Introduction

Malnutrition poses significant health risks by impairing essential physiological functions, including growth, maintenance, and energy metabolism. These risks are particularly pronounced during pregnancy and lactation, periods marked by elevated nutritional demands. During these periods, deficiencies of essential nutrients, such as iodine, iron, folate, calcium, and zinc, have been associated with an increased incidence of anemia, pre-eclampsia and eclampsia, hemorrhage, stillbirth, low birth weight, intrauterine growth restriction, and neural tube defects (1,2).

Globally, over 1 billion adolescent girls and women suffer from malnutrition. In Africa, up to 23.5% of pregnant women are believed to be malnourished (3). Following the recent global food and nutrition crisis, the number of acutely malnourished pregnant and lactating women in 12 low income countries, including Ethiopia, has also increased by 25% (4).

Malnutrition may arise not only from inadequate nutrient intake, malabsorption, and metabolic dysfunction but also from infectious comorbidities, such as HIV/AIDS. The interplay between HIV/AIDS and malnutrition is synergistic and can lead to profound adverse health outcomes, particularly during pregnancy (5–7). HIV/AIDS may exacerbate pre-existing malnutrition or independently precipitate nutritional deficiencies by reducing nutrient intake through opportunistic infection–related anorexia and dysphagia, impairing gastrointestinal absorption, and elevating the body’s metabolic demands (5–7). The resultant malnutrition, in turn, contributes to disease progression by diminishing the immune system’s capacity to combat infection (5–8).

Since its emergence in the 1980s, HIV/AIDS has claimed the lives of more than 44 million people and continues to pose a significant public health challenge worldwide (9,10). Current estimates indicate that more than 40 million people are infected by the virus, of whom 53% are women and girls (9,11). The total number of pregnant women living with the virus is also estimated at 1.1 million, with the sub-Saharan region disproportionately bearing 90% of the disease burden (9,11,12).

Human immuno deficiency virus can be transmitted through infected blood, semen, and vaginal secretions and may also be passed from mother to child in utero during delivery or breastfeeding (13). According to the World Health Organization, without intervention, between 15% and 45% of infants born to HIV-positive mothers acquire the virus during pregnancy, delivery, or breastfeeding (14). Maternal malnutrition further intensify this risk, increasing the likelihood of mother-to-child transmission (MTCT) by up to twofold (15,16). Malnutrition also elevates the risk of adverse obstetric outcomes, including preterm birth, stillbirth, and low birth weight, particularly among women living with HIV (8,15,16).

To mitigate the risk of MTCT and accelerate the progress toward the UN goal of ending AIDS as a public health threat by 2030, the Global Plan toward the Elimination of New HIV Infections among Children and Keeping their Mothers Alive was launched in 2011. Subsequently, an estimated 2.1 million deaths and 4.4 million HIV infections among pregnant women and children have been averted (17,18). This success is largely attributable to the expansion of prevention of mother-to-child transmission (PMTCT) services and increased initiation of lifelong antiretroviral therapy for pregnant women living with the virus (17). Despite this, however, 120,000 new pediatric HIV infections were recorded in 2024, putting at risk the progress made to attain the 2030 targets (17).

Ethiopia has achieved notable progress in combating the HIV epidemic over the past decade, reflected in a decline in national adult (15–49) HIV prevalence from 1.97% in 1990 to 0.93% in 2019 (19). Treatment coverage has also expanded, with 90% of people living with HIV aware of their status, 94% on antiretroviral therapy, and 96% achieving viral suppression by 2025 (19). Mother-to-child transmission has also decreased markedly, from 39.55% in 2000 to 16.9% in 2019, with accelerated improvements following the introduction of the B+ option in 2013 (12,19). Building on these achievements, ambitious targets, aligned with the global 95-95-95 goals were integrated into the Health Sector Transformation Plan II and the National Strategic Plan for HIV (2021–2025). These plans commit to reducing mother-to-child transmission to less than 5% by 2025 and below 2% by 2030 (19,20). To achieve these targets, nutrition support was incorporated into the care of pregnant and lactating women living with HIV under the National Nutrition Program (NNP) and the National Nutrition and HIV/AIDS Implementation (21,22). These programs prioritize tailored dietary counseling, supplementation, and food security measures to enhance maternal and child health (21,22). However, these advances have not yet translated into sufficient impact, as the mother-to-child transmission (MTCT) rate in Ethiopia remains unacceptably high. Furthermore, a significant contributing factor is the high prevalence of malnutrition, affecting up to 43% of HIV-positive pregnant and lactating women mostly due to household food insecurity, inadequate dietary diversity, anemia, and absence of nutritional support (23–25).

In conclusion, although malnutrition severely threatens the health of HIV-positive mothers and efficacy of PMTCT programs, research on its extent and causes in this vulnerable group remains inadequate. The existing evidence on prevalence remains variable and region-specific, while reported determinants, including socioeconomic, clinical, and treatment-related factors, remain inconsistent. A focused investigation within the dense and rapidly evolving urban context of Addis Ababa is notably absent. This need is especially urgent given Ethiopia’s recent and substantial decrement in foreign funding, which historically accounted for up to 53% of its health sector financing. This shift heightens the vulnerability of essential programs, including PMTCT, and necessitates precise, locally-grounded evidence to ensure efficient and sustainable allocation of diminished resources. This study addresses this gap by systematically assessing the prevalence and key determinants of malnutrition among HIV-positive pregnant and lactating women receiving PMTCT care in Addis Ababa’s public hospitals. The findings will provide essential, localized evidence to inform targeted nutritional interventions and refine integrated HIV care strategies, ensuring they are both resilient and cost effective within the current fiscal landscape. This will directly contribute to improved health outcomes for this vulnerable population and their children.

## Methods and Materials

### Study design, area, and period

This institution-based cross-sectional study, aimed at assessing the prevalence of malnutrition and associated factors among HIV-positive pregnant and lactating women receiving PMTCT care, was conducted from April 1 to April 20, 2024, in public hospitals located across the 11 sub-cities of Addis Ababa, Ethiopia. Addis Ababa, the nation’s capital and largest urban center, is home to an estimated population of 3.38 million people and serves as a major hub for specialized healthcare services, including comprehensive PMTCT programs.

### Populations

The source population for this study consisted of all pregnant and lactating women living with HIV who were actively enrolled in and receiving PMTCT care at public hospitals within the city. The study population was derived from this source and comprised pregnant and lactating women living with HIV who were attending PMTCT clinics at public hospitals in Addis Ababa.

### Sample size determination

A single population proportion formula was used to calculate the minimum sample of participants required to accurately estimate the outcome of interest. The prevalence of malnutrition among pregnant and lactating women living with HIV/AIDS was estimated to be 34.2% based on a prior study (25). A 95% confidence interval (Z = 1.96) and a 5% margin of error were also considered. As the total population of pregnant and lactating women receiving PMTCT care in the selected hospitals was 374, a finite population correction (FPC) was applied. After the sample size was corrected and a 15% nonresponse rate was added, the final sample size became 212.

### Sampling technique and procedure

A random sample of six public hospitals was selected from the 11 in Addis Ababa. From the PMTCT client lists of these hospitals, the study participants were selected via systematic random sampling. Using a calculated sampling interval of approximately 2, every second client was chosen. In cases where a selected client was ineligible, the next client on the list was selected sequentially until the predetermined sample size was met.

### Data collection method

Data were collected via interviewer-administered questionnaires, anthropometric measurements (BMI, MUAC), and medical record review. The questionnaires covered sociodemographic, clinical, and dietary diversity variables. Clinical data on hemoglobin, CD4 count, ART adherence, and opportunistic infections were extracted from patient files. Anthropometric measurements followed standard protocols, with inter-observer technical error of measurement calculated to ensure precision.

Rigorous quality control protocols were implemented to ensure data integrity. These included pre-testing the questionnaire at a non-participating facility one week prior to data collection, which led to refinements in the clarity of several items. All data collectors and supervisors underwent a one-day standardized training on ethical interview techniques, accurate anthropometric measurement, and unbiased data extraction from medical records to ensure procedural consistency. The assigned supervisors also conducted daily reviews to validate data completeness and logical consistency.

### Data processing and analysis

The data were loaded into SPSS version 26, after which descriptive statistical analysis was conducted to generate summary figures in the form of frequencies and percentages. Furthermore, bivariable and multivariable logistic regression models were applied to investigate the relationships between the the outcome variable and explanatory variables. During the bivariable analysis, a p value ≤ 0.25 was used as a criterion for including variables into the multivariable model, whereas in the multivariable model, a p value less than 0.05 was used as an indicator of statistical significance.

### Ethical considerations

The study received ethical clearance from the Institutional Review Board of Asrat Woldeyes Health Science Campus, Debre Birhan University, and permission from the Addis Ababa Health Bureau. All participants provided written informed consent following a verbal explanation. Data were anonymized to ensure confidentiality.

## Results

### Sociodemographic characteristics of the respondents

A total of 212 eligible women were approached, of whom 193 participated, yielding a response rate of 91%. The participants were predominantly aged 25–34 years (47.2%) or 35–49 years (45.1%). Regarding education, the largest proportions had no formal education (31.1%) or primary education (28.0%). Most participants were also married (40.9%) or widowed (22.3%). The most common occupations were government employment (27.5%) and private sector employment (25.4%). More than half of the respondents (60.1%) reported a monthly income below 2500 ETB, and the majority (79.8%) lived in households with four or fewer members (**Table 1).**

**Table 1:** Sociodemographic characteristics of pregnant and lactating women living with HIV/AIDS receiving PMTCT care in public hospitals in Addis Ababa, Ethiopia, 2024.

| **Variables** | **Categories** | **Malnourished (%)** | **Non malnourished (%)** | **Total (%)** |
| --- | --- | --- | --- | --- |
| **Age** | 15-24 years | 3 (1.6%) | 12 (6.2%) | 15 (7.8%) |
|  | 25-34 years | 17 (8.8%) | 74 (38.3%) | 91 (47.2%) |
|  | 35-49 years | 36 (18.7%) | 51 (26.4%) | 87 (45.1%) |
| **Educational status** | No formal education | 20 (10.4%) | 40 (20.7%) | 60 (31.1%) |
|  | Primary school | 19 (9.8%) | 35 (18.1%) | 54 (28.0%) |
|  | Secondary school | 13 (6.7%) | 27 (14.0%) | 40 (20.7%) |
|  | University/College | 4 (2.1%) | 35 (18.1%) | 39 (20.2%) |
| **Marital status** | Single | 7 (3.6%) | 32 (16.6%) | 39 (20.2%) |
|  | Married | 25 (13.0%) | 54 (28.0%) | 79 (40.9%) |
|  | Divorced | 15 (7.8%) | 17 (8.8%) | 32 (16.6%) |
|  | Widowed | 9 (4.7%) | 34 (17.6%) | 43 (22.3%) |
| **Occupation** | Government Employee | 11 (5.7%) | 42 (21.8%) | 53 (27.5%) |
|  | Daily Laborer | 5 (2.6%) | 12 (6.2%) | 17 (8.8%) |
|  | Housewife | 5 (2.6%) | 18 (9.3%) | 23 (11.9%) |
|  | Private-Employee | 12 (6.2%) | 37 (19.2%) | 49 (25.4%) |
|  | Unemployed | 23 (11.9%) | 28 (14.5%) | 51 (26.4%) |
| **Income** | < 2500 ETB | 43 (22.3%) | 73 (37.8%) | 116 (60.1%) |
|  | ≥2500 ETB | 13 (6.7%) | 64 (33.3%) | 77 (39.9%) |
| **Family Size** | <5 | 49 (25.4%) | 105 (54.4%) | 154 (79.8%) |
|  | 5-10 | 7 (3.6%) | 32 (16.6%) | 39 (20.2%) |

### Clinical profile of the respondents

Nearly half (42.5%) of the respondents had been receiving antiretroviral therapy (ART) for over a decade. A minority (15.5%) reported experiencing side effects from ART, whereas the vast majority (84%) had never encountered any. Gastrointestinal symptoms and eating problems were reported by 34.7% and 18% of the participants, respectively. Regarding reproductive health, most participants (60%) had given birth to two or fewer children, and 13.5% were pregnant at the time of the study.

Clinically, a serum hemoglobin level below 12 g/dL was observed in 22.3% of respondents. More than half (58%) had a CD4+ count exceeding 500 cells/mm³, and nearly all participants (92.2%) were classified in WHO clinical stages 1 or 2. Adherence to ART was good for 50% of respondents, whereas treatment failure was recorded for 11.4%. Concerning co-infections, Hepatitis C Virus (HCV) and Hepatitis B Virus (HBV) were detected in 20% of the study participants. However, no history of syphilis infection was found in any of the participants. A recent viral load exceeding 1000 copies/ml was recorded for 16 respondents (8.3%). The majority of mothers (60.6%) also received cotrimoxazole preventive therapy (CPT) at one point during their visit (**Table 2**).

**Table 2:**
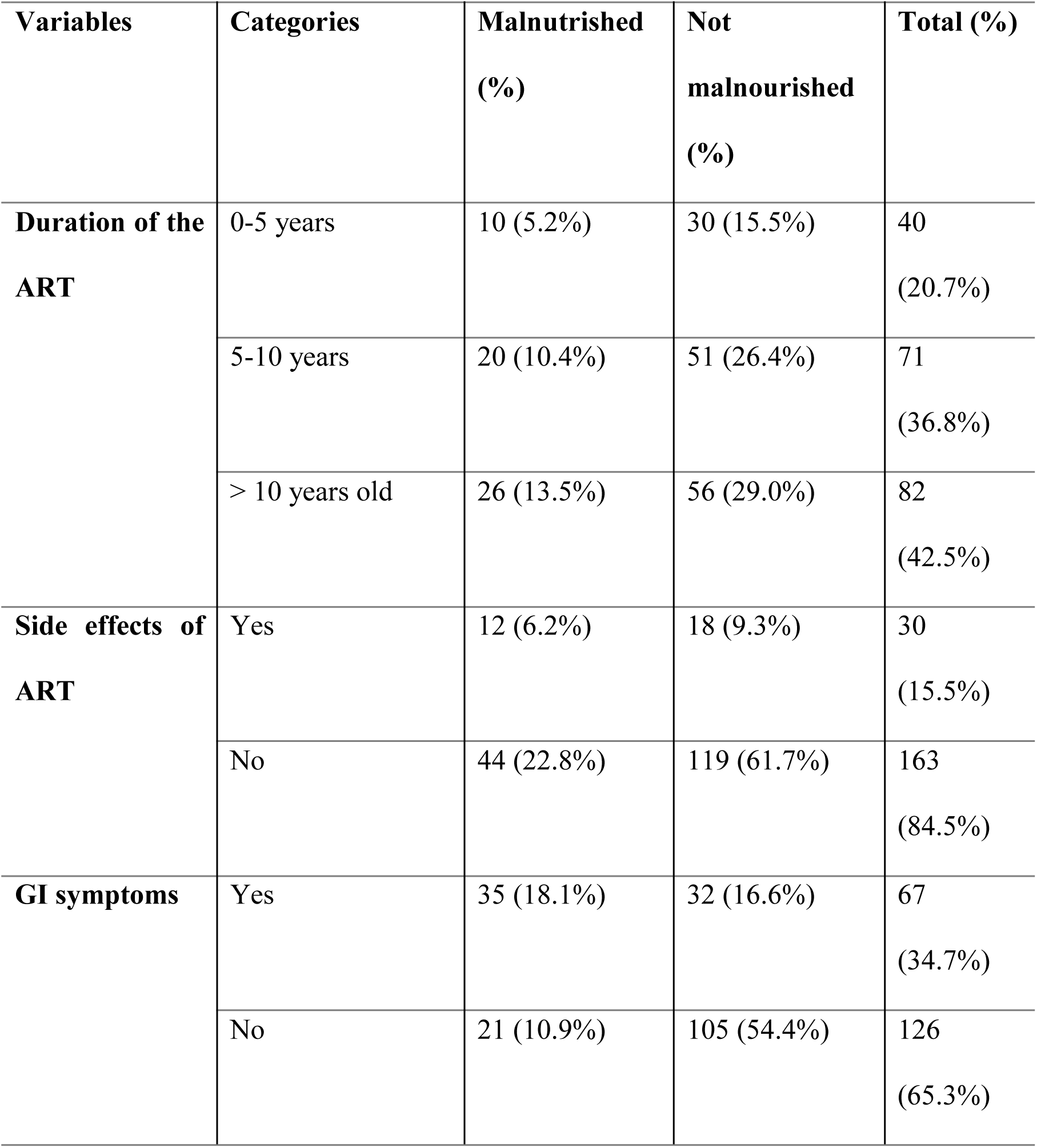
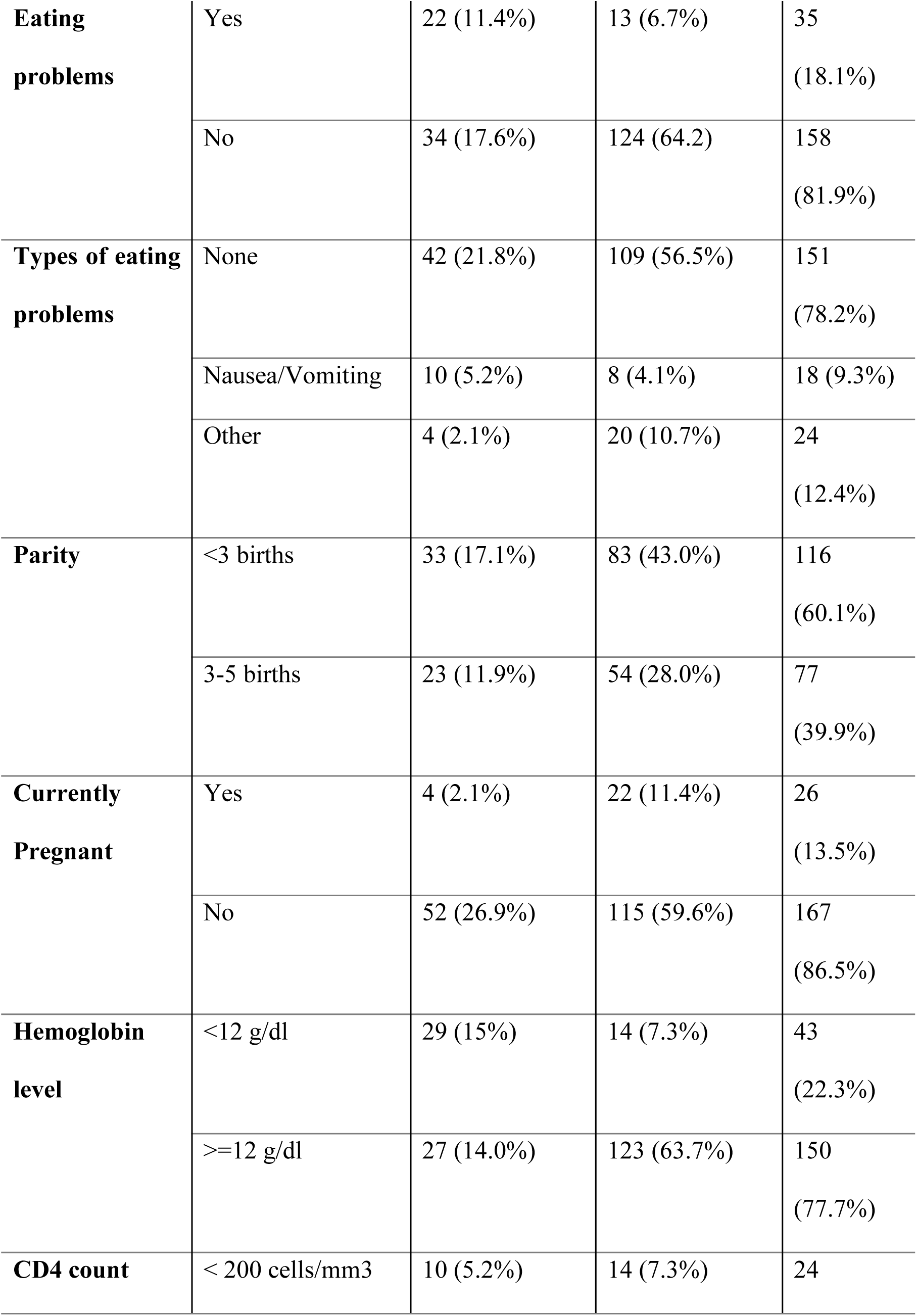
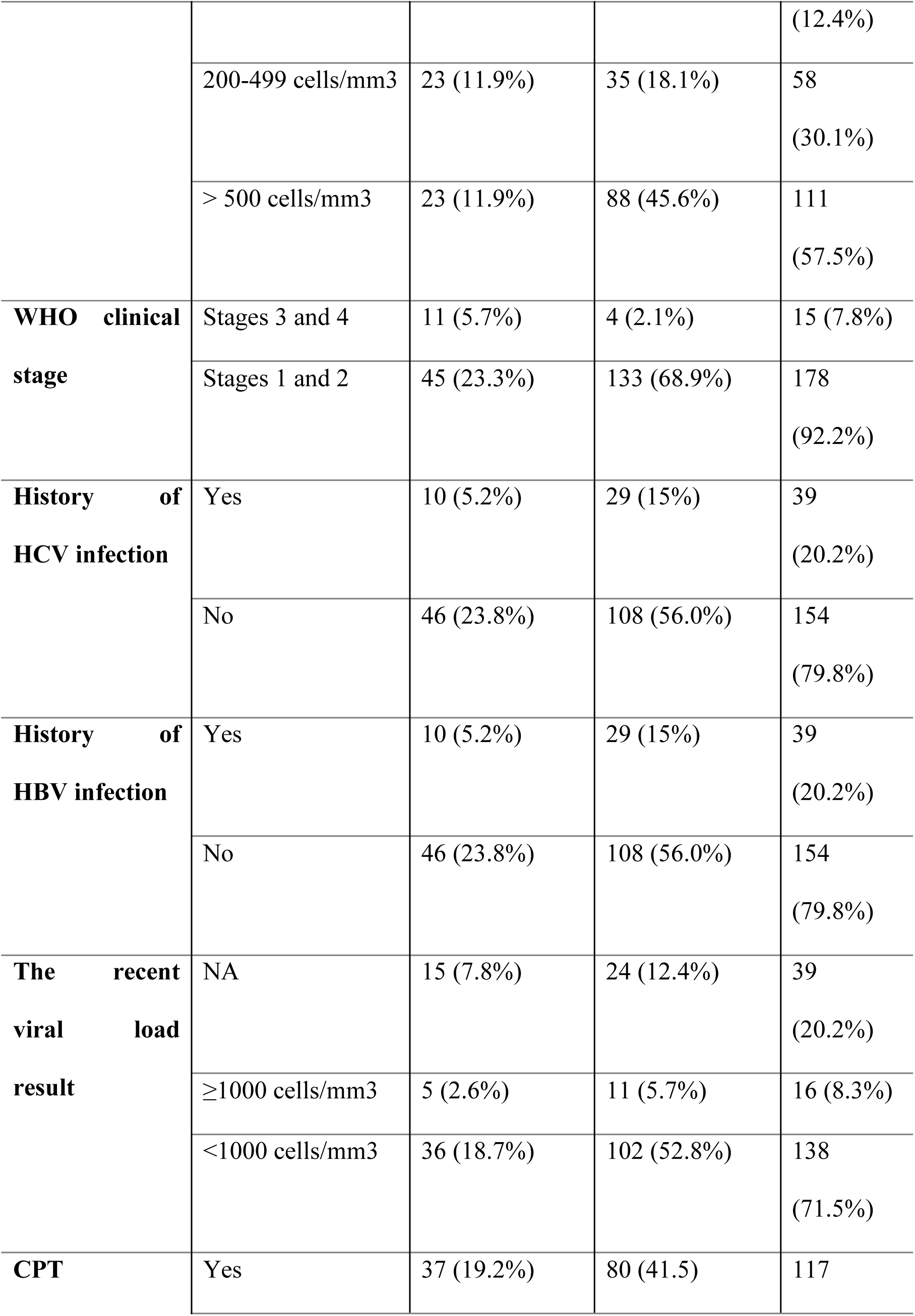
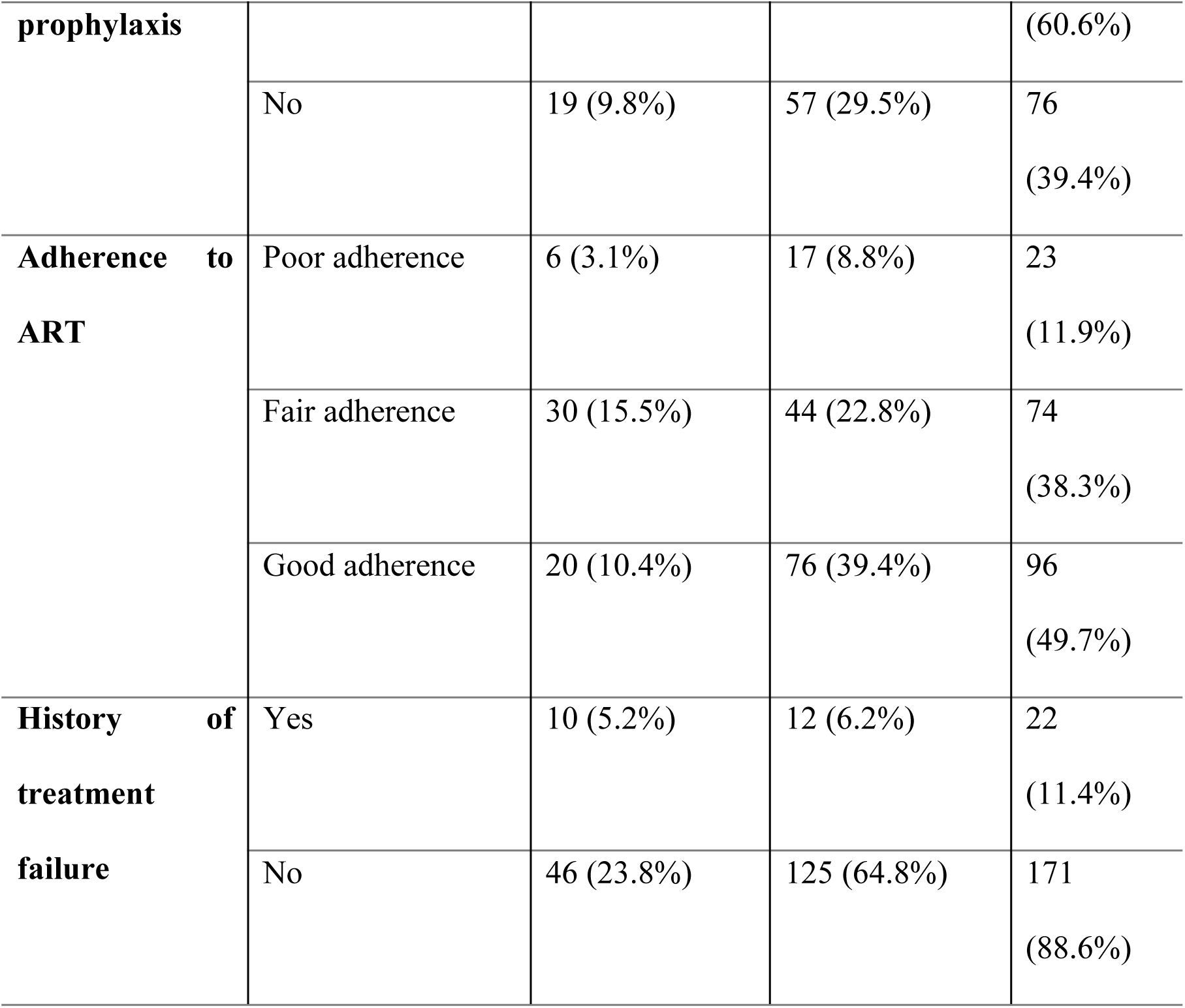
Clinical and health-related characteristics of pregnant and lactating women living with HIV/AIDS receiving PMTCT care at public hospitals in Addis Ababa, Ethiopia, 2024.

### Dietary practice of the respondents

Approximately one-quarter of participants (21.8%) reported altering their dietary habits. Of them, 16.6% modified both the quality and quantity of their food, whereas 12.4% changed their meal frequency. Overall, nearly two-thirds of respondents (62.7%) consumed more than three meals per day. More than half (60.1%) of the participants had also received dietary consultation. However, only 37.3% met the minimum dietary diversity. Regarding the food consumption pattern of the respondents, 23.8% had a poor food consumption score (**Table 3**).

**Table 3:** Dietary-related practices of pregnant and lactating women living with HIV/AIDS receiving PMTCT care in public Hospitals of Addis Ababa, Ethiopia, 2024.

| **Variables** | **Categories** | **Malnourished (%)** | **Not malnourished (%)** | **Total (%)** |
| --- | --- | --- | --- | --- |
| **Changed feeding style** | Yes | 14 (7.3%) | 28 (14.5%) | 42 (21.8%) |
|  | No | 42 (21.8%) | 109 (56.5%) | 151 (78.2%) |
| **Type of feeding style change** | Not Applicable | 42 (21.8%) | 109 (56.5%) | 151 (78.2%) |
|  | Frequency | 4 (2.1%) | 7 (4.1%) | 11 (6.2%) |
|  | Food quality and quantity | 10 (5.2%) | 21 (11.4%) | 31 (15.6%) |
| **Meal frequency** | <3 meals | 38 (19.7%) | 34 (17.6%) | 72 (37.3%) |
|  | Three or more meals | 18 (9.3%) | 103 (53.4%) | 121 (62.7%) |
| **Received dietary counseling** | No | 22 (11.4%) | 55 (28.5%) | 77 (39.9%) |
|  | Yes | 34 (17.6%) | 82 (42.5%) | 116 (60.1%) |
| **DDS** | Low DDS | 23 (11.9%) | 34 (17.6%) | 57 (29.5%) |
|  | Medium DDS | 21 (10.9%) | 43 (22.3%) | 64 (33.2%) |
|  | High DDS | 12 (6.2%) | 60 (31.1%) | 72 (37.3%) |
| **FCS** | Poor FCS | 27 (14%) | 19 (9.8%) | 46 (23.8%) |
|  | Borderline FCS | 17 (8.8%) | 72 (37.3%) | 89 (46.1%) |
|  | Acceptable FCS | 12 (6.2%) | 46 (23.8%) | 58 (30.1%) |

### Prevalence of malnutrition

The overall prevalence of malnutrition among pregnant and lactating women living with HIV was 29%. It was notably higher among lactating (non-pregnant) mothers (31%) than pregnant mothers (15.4%). The prevalence of undernutrition, overweight, and obesity was 19%, 8%, and 2%, respectively (**Fig. 1**).

**Fig 1:**
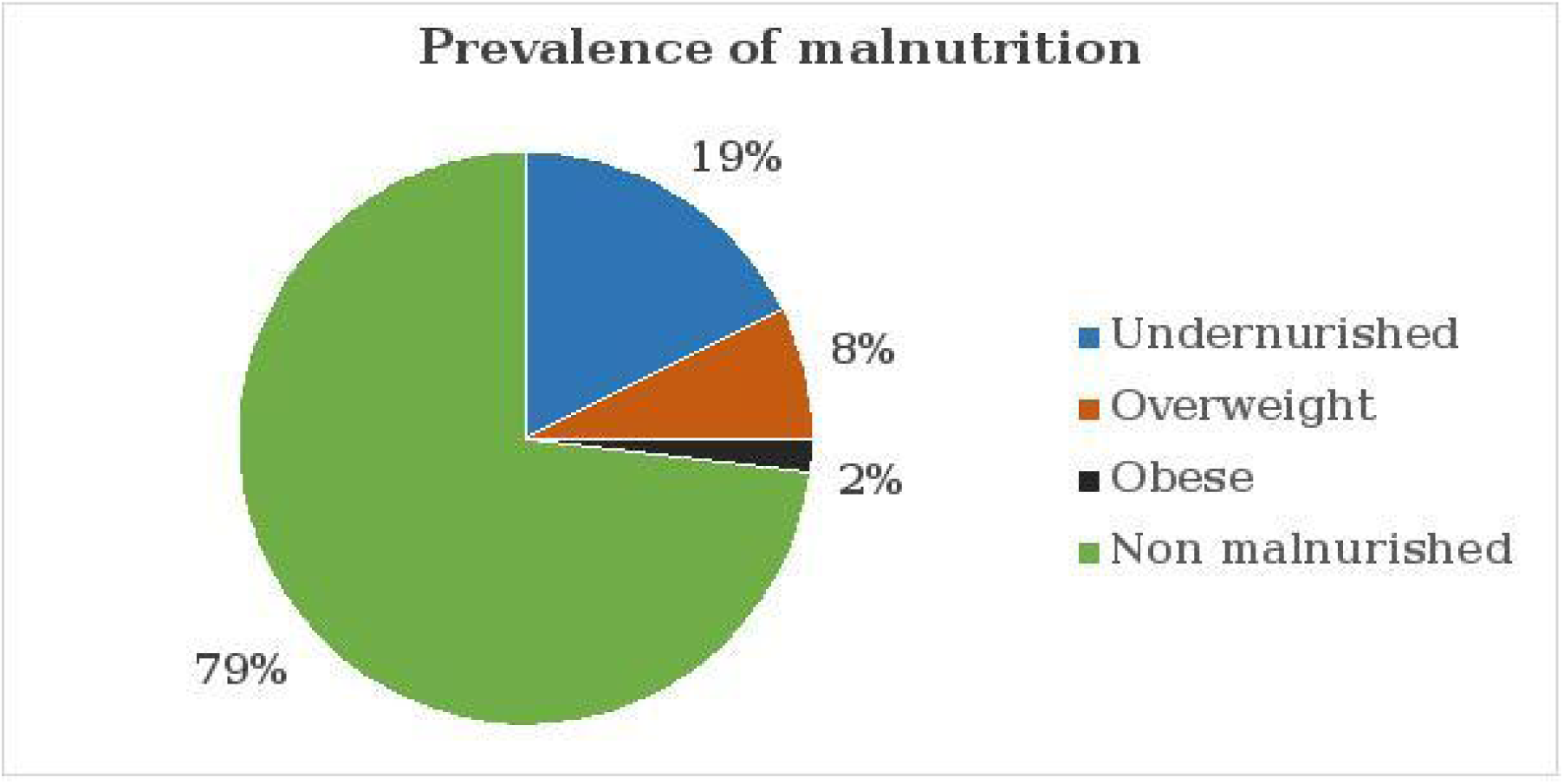
Prevalence of malnutrition among pregnant and lactating women living with HIV/AIDS receiving PMTCT care at public hospitals in Addis Ababa, Ethiopia, 2024.

### Factors associated with malnutrition

Binary logistic regression was used to determine the associations between predictor variables and the nutritional status of pregnant and lactating women living with HIV/AIDS and receiving PMTCT care. Initially, each independent variable was entered into a bivariable logistic regression. Variables with p value scores less than 0.25 were selected for further analysis in multivariable logistic regression model. These variables included age, educational status, occupation, income, family size, meal frequency pattern, dietary counseling, duration of ART, eating problems, gastrointestinal symptoms, hemoglobin level, CD4 cell count, WHO clinical staging, viral load result, adherence to ART, treatment failure, dietary diversity score (DDS), and food consumption score (FCS).

According to the multivariable logistic regression analysis, age, meal frequency, gastrointestinal symptoms, eating problems, hemoglobin level, CD4 count, and adherence to ART were significantly associated with malnutrition (p value < 0.05).

Specifically, HIV-infected pregnant and lactating women aged 25-34 years had 76% lower odds of malnutrition than those aged 35-49 years (AOR: 0.248). In contrast, those who ate fewer than three meals per day had 3.2 times higher odds of malnutrition than those who ate three or more meals daily (AOR: 4.12). Additionally, HIV-infected pregnant and lactating women with gastrointestinal (GI) symptoms had 3.5 times higher odds of malnutrition than those without such symptoms (AOR: 3.52). Eating problems posed a particularly high risk, with HIV-infected pregnant and lactating women reporting eating issues having 13 times more odds of malnutrition than those without eating problems (AOR: 13.7). Low hemoglobin levels (below 12 mg/dl) in HIV-infected pregnant and lactating women have also increased the odds of malnutrition significantly, up to 5 times (AOR: 5.03). Immunosuppression in HIV-infected pregnant and lactating women, as indicated by a CD4 count of less than 200 cells/mm³, further increases the odds of malnutrition by 8 factors (AOR: 8.13). Finally, poor adherence to treatment was a strong risk factor, as those with poor adherence having 3.6 times higher the odds of malnutrition than those with good adherence (AOR: 3.6) (**Table 4**).

**Table 4:** Binary logistic regression analysis of factors associated with malnutrition among pregnant and lactating women living with HIV/AIDS receiving PMTCT care in public hospitals in Addis Ababa, Ethiopia, 2024.

| Variables | Categories | Malnourished (%) | Non-malnourished (%) | AOR | 95% C.I. | p-value |
| --- | --- | --- | --- | --- | --- | --- |
| **Age** | 15-24 years | 2 (1.0%) | 13 (6.7%) | 0.996 | 0.226 - 4.398 | 0.996 |
|  | 25-34 years | 17 (8.8%) | 73 (37.8%) | 0.248 | 0.094 - 0.651 | 0.005 |
|  | 35-49 years | 37 (19.2%) | 51 (26.4%) | 1 | 1 | 1 |
| **Meal frequency** | Three or more meals | 18 (9.3%) | 103 (53.4%) | 1 | 1 | 1 |
|  | <3 meals | 38 (19.7%) | 34 (17.6%) | 4.115 | 1.585 - 10.688 | 0.004 |
| **GI symptoms** | No GI symptoms | 51 (26.4%) | 104 (53.9%) | 1 | 1 | 1 |
|  | Yes (GI symptoms) | 5 (2.6%) | 33 (17.1%) | 3.518 | 1.355 - 9.136 | 0.010 |
| **Eating problems** | No Eating Problem | 26 (13.5%) | 123 (63.7%) | 1 | 1 | 1 |
|  | Yes (Eating Problem) | 30 (15.5%) | 14 (7.3%) | 13.7 | 3.85 - 47.62 | 0.004 |
| **Hemoglobin levels** | ≥ 12 mg/dL | 27 (14.0%) | 123 (63.7%) | 1 | 1 | 1 |
|  | <12 mg/dl | 29 (15.0%) | 14 (7.3%) | 5.032 | 1.46 - 17.24 | 0.001 |
| **CD4 count** | >500 cells/mm<sup>3</sup> | 26 (13.5%) | 86 (44.6%) | 1 | 1 | 1 |
|  | 200-499 cells/mm<sup>3</sup> | 20 (10.4%) | 37 (19.2%) | 1.02 | 0.36 - 2.87 | 0.966 |
|  | <200 cells/mm<sup>3</sup> | 10 (5.2%) | 14 (7.3%) | 8.131 | 1.37 - 47.62 | 0.019 |
| **Adherence to ART** | Good adherence | 10 (5.2%) | 78 (40.4%) | 1 | 1 | 1 |
|  | Fair adherence | 14 (7.3%) | 32 (16.6%) | 2.599 | 0.876 - 7.705 | 0.085 |
|  | Poor adherence | 32 (16.6%) | 27 (14.0%) | 3.625 | 1.078 - 40.710 | 0.041 |

## Discussion

This study aimed to assess the prevalence of malnutrition and associated factors among pregnant and lactating women living with HIV/AIDS receiving PMTCT care in public hospitals of Addis Ababa. A total of 193 pregnant and lactating women living with HIV/AIDS were included in this study.

The overall prevalence of malnutrition among this cohort was 29%, comprising undernutrition (19%), overweight (8%), and obesity (2%). The rate of undernutrition (19%) aligns with findings from other Ethiopian and non-Ethiopian studies, including those conducted in Asella (18.3%), Legatafo (18.8%), Shashemene (15.9%), and Arba Minch (18.2%), Rwanda (22%), and Nepal (18.3%) (26–31). However, it is notably lower than the 29.2% prevalence reported in the Bench Sheko Zone and higher than the 10.3% reported in Bushenyi district in southwestern Uganda (32,33). While comparing the prevalence of overnutrition obtained from our study (overweight 8%, obesity 2%) with findings from Kenya (overweight 17.2%, obesity 3.6%) and Southwest Ethiopia (overweight 9.6%), our rates are notably lower than those reported in Kenya but similar to the overweight prevalence in Southwest Ethiopia (33,34). These variations might be explained by differences in local food security, agricultural conditions, and the coverage of health interventions, compounded by this cohort’s distinct clinical profile as HIV-positive women and variable nutritional integration within local PMTCT services. Concurrently, the lower prevalence of overnutrition compared to Kenya may reflect Kenya’s more advanced stage of the nutrition transition, driven by greater urbanization, whereas similarities with other Ethiopian studies underscore shared regional socioeconomic and lifestyle patterns.

According to the multivariate logistic regression analysis, age, meal frequency, GI symptoms, eating problems, hemoglobin level, CD4 count, and adherence to ART were significantly associated with malnutrition in the study area.

This study identified advanced age as a significant predictor of malnutrition among HIV-infected pregnant and lactating women, with older individuals exhibiting a higher risk than their younger counterparts. This finding aligns with research conducted in the Maji Zone of Southwest Ethiopia, which similarly concluded that older HIV-infected adults are more susceptible to malnutrition than younger adults (35). However, this result contrasts with a substantial body of literature indicating that younger age is associated with an increased risk of malnutrition among people living with HIV (31,33,36–40). Nevertheless, our finding can be explained by the fact that older individuals often experience a natural weakening of the immune system, which can be accelerated by HIV progression. This immunosenescence may impair the body’s ability to combat infections and maintain metabolic balance. Consequently, older women are likely more susceptible to opportunistic infections (e.g., oral candidiasis, chronic diarrhea) that directly compromise nutritional intake, absorption, and utilization. In contrast, the typically more robust immune function in younger women may provide greater resilience against such comorbidities, offering some protection against rapid nutritional deterioration during pregnancy and lactation.

In this study, meal frequency was also identified as a significant factor associated with malnutrition. Women who consumed fewer than three meals per day were four times more likely to be malnourished than those who ate three or more times within a 24-hour period. This finding is consistent with evidence from multiple settings including those conducted in Hosana and Shashemene, as well as with a study in Tanzania, which similarly reported that individuals eating fewer meals daily were approximately more likely to develop malnutrition (26,41,42). The biological plausibility of this association is strong, particularly for HIV-infected pregnant and lactating women, who have substantially elevated nutritional demands. Frequent meal consumption helps maintain positive nitrogen balance, stabilizes energy availability, and supports the preservation of lean body mass. This consistent nutrient intake is critical for sustaining immune competence, which may enhance resilience against opportunistic infections, such as oral candidiasis or persistent gastroenteritis, that directly impair nutrient absorption and utilization, thereby worsening malnutrition. Consequently, inadequate meal frequency likely creates a cycle of depleted energy reserves and compromised immunity, accelerating nutritional decline in this vulnerable population.Meal frequency is another variable significantly associated with malnutrition.

The study further showed that respondents with gastrointestinal symptoms were more likely to be malnourished than their counterparts were. This finding was similar to that of a study conducted in Rwanda, which showed that having GI symptoms significantly increases the risk of malnutrition (30). The explanation for this could be that GI discomfort, with or without nausea and vomiting, may significantly reduce the frequency and amount of meals, as well as the amount of nutrients absorbed from the GI tract of HIV-infected pregnant and lactating women, reducing their ability to fight infection and hastening the progression of the disease to full-blown AIDS. Furthermore, experiencing significant GI discomfort may make it difficult for HIV-infected pregnant and breastfeeding women to adhere to their ART regimen. These factors, together with the high energy demand state of pregnancy and nursing, experiencing GI symptoms among HIV-infected pregnant and lactating women could considerably expose them to malnutrition (6,43). For similar reasons, pregnant and lactating women living with HIV/AIDS who had difficulty eating food might develop malnutrition (44).

In our study, low hemoglobin levels were also found to significantly affect the nutritional status of pregnant and lactating women on ART. In line with our findings, studies conducted in Nepal and Ethiopia revealed that having low Hb levels increased the odds of malnutrition (31,45). This finding likely stems from the high prevalence of iron deficiency anemia observed in reproductive-age women in Ethiopia, even before pregnancy. This condition is further exacerbated by the increased physiological iron demands that arise during and after pregnancy. Additionally, the chronic nature of HIV infection may further deplete the body’s iron stores, potentially exposing HIV-infected pregnant women to a higher risk of anemia.

This study also indicated that the CD4 count was a significant factor associated with malnutrition. This was supported by studies from Senegal, Tanzania, the Bench-Sheko zone, and Shashemene, which showed a significant association between low CD4 counts and malnutrition (26,33,46,47). These findings amplify the relationship between immune suppression and nutritional status in HIV patients. A low CD4+ T-cell count suggests advanced HIV progression and compromised immunity, increasing vulnerability to opportunistic infections that decrease nutrient absorption and increase energy demands. This generates a vicious cycle in which poor immune function exacerbates starvation, which in turn decreases immunological responses and increases the risk developing malnutrition.

In addition, ART adherence was found to affect the nutritional status of HIV infected pregnant and lactating women. Pregnant and lactating women who had poor ART adherence had greater odds of developing malnutrition than did those who had good ART adherence. These findings are supported by studies performed in the Bench-Sheko zone and Addis Ababa, which revealed that patients who had good ART adherence were less likely to develop malnutrition (33,48). Similarly, a study conducted in Uganda revealed that clients with poor adherence are more likely to be malnourished than those with good adherence (32). This may be explained by the fact that poor ART adherence accelerates viral replication, leads to the destruction of CD4+ T cells, weakens the immune system, and supercharges disease progression. As a result, this can expose patients to opportunistic infections, ultimately leading to reduced dietary intake and impaired nutrient absorption.

## Conclusions and recommendations

This study showed that the prevalence of malnutrition among women receiving PMTCT care at public hospitals in Addis Ababa was high. Age, meal frequency, GI symptoms, eating problems, hemoglobin levels, CD4 counts, and adherence to ART were also significantly associated with malnutrition among pregnant and lactating women living with HIV/AIDS receiving PMTCT care in Addis Ababa public hospitals.

Moving forward, integrating routine nutritional screening and evidence-based supportive interventions, including dietary support for women with low meal frequency and targeted management for those with anemia or low CD4 counts, should be a priority to improve comprehensive care for this vulnerable population.

## Data Availability

The datasets used and/or analyzed during the current study are available from the corresponding author upon reasonable request.

## Declarations

### Consent for publication

Not applicable

